# Caregiver ECHO: A Model for Delivering Virtual Behavior Management Education to Families of Children with Developmental Disabilities

**DOI:** 10.1101/2024.02.10.24302623

**Authors:** Rose E. Nevill, Gail E. Lovette, Katherine J. Bateman, Natalie M. Badgett, Genevieve R. Lyons, Emily C. Fuhrmann, Genevieve E. Bohac, Karen G. Orlando, Keith D. Page

## Abstract

Children with developmental disabilities are at high risk of challenging behavior, yet their families experience consistent barriers to affordable parent education in behavior management. The present study tested the efficacy of a caregiver-focused Extensions of Community Health Outcomes program for delivering virtual parent education and support to familial caregivers of children with DD in behavior management. A pre-post quasi-experimental design was used to evaluate the effect of Caregiver ECHO on caregiver participants’ (n = 30) knowledge of behavior modification strategies, self-efficacy in managing challenging behavior, empowerment, and negative emotional reactions to challenging behavior. Participating in Caregiver ECHO resulted in significant increases in caregiver knowledge, self-efficacy, and empowerment, and significant reductions in certain negative emotional reactions to challenging behavior. The Caregiver ECHO model offers advantages over other parent education programs in that it emphasizes peer learning, active problem-solving, and community building as core components of its approach while using low-cost methodologies.

## Caregiver ECHO: A Model for Delivering Virtual Behavior Management Education to Families of Children with Developmental Disabilities

Children with developmental disabilities (DD) display higher rates of challenging behavior (CB) than children without DD (Emerson et al., 2014; Nicholls et al., 2020). While evidence-based models for addressing CB have been developed, such as applied behavior analysis and positive behavior supports, these treatments remain difficult for families of children with DD to access. Many states only mandate insurance coverage of behavioral therapy for people with an autism spectrum disorder (ASD) diagnosis, despite ample evidence of behavioral principles being effective across behaviors and diagnoses (Heinicke & Carr, 2014; LaRue et al., 2015; Salloum et al., 2016). Affordable access to behavioral therapies also remains challenging due to the limited number of qualified providers, long waitlists, age discrimination, and inequity in diagnostic access across racial and ethnic minority groups (Trump & Ayres, 2019). Lacking access to behavioral supports risks increasing the vulnerabilities that many families of children with DD already face: social isolation (Halstead et al., 2018), parenting stress (Barroso et al., 2018), and overall poor quality of life (Zeng et al., 2020). As such, further research identifying inexpensive and accessible methods for delivering behavior supports to a broad range of DD is needed (Zeng et al., 2020).

Parent education in evidence-based interventions has been established as an effective method for increasing families’ capacities to address child CB. Parent education in behavioral interventions, specifically, has been shown to effectively reduce CB (Machalicek et al., 2016; Pennefather et al., 2018; Vismara et al., 2018) and increase parents’ self-efficacy in behavior management (Bearss et al., 2018; Lequia et al., 2013). One key benefit of parent education programs is that they are typically delivered in a group format to parents of children with similar behavioral profiles or diagnoses, thereby not only providing valuable education but also access to a social network of like-minded peers. Indeed, social support has been identified as a key predictor of resilience among parents of children with DD (Peer & Hillman, 2014).

### The ECHO Model

The Extensions of Community Health Outcomes (ECHO) model is an innovative telehealth-based model that provides education and opportunities for community building (Arora, 2015) – a feature that is particularly relevant for addressing the typical social isolation experienced by families of children with DD. The four core ingredients of the ECHO model are a) using technology to leverage scarce resources, b) disseminating best practices to reduce disparities, c) using case-based learning to build expertise, and d) outcomes monitoring through data collection (Project ECHO, 2021). ECHOs virtually connect an interdisciplinary “hub” team of experts, typically university-based clinicians, with community-based practitioners or “spokes”, over a free teleconferencing platform such as Zoom® for regularly scheduled ECHO sessions to provide education and support around a condition (e.g., autism) or problem (e.g., burnout amongst healthcare workers). In each session, a hub team member delivers a short 20– 25-minute workshop on a relevant topic, and a community-based spoke presents the case of a deidentified patient or client with a guiding question (e.g., “should my patient be evaluated for ASD?”) for peer consultation. The deidentification of patients is important for facilitating cross-state and cross-region collaboration between and consultation from licensed healthcare professionals. The hub and spokes then engage in group-based problem solving to generate recommendations for the spoke. This interactive learning approach has been shown to enable all participants to learn from expert hub members as well as their peers, and in the process build a supportive community (Arora et al., 2011).

Previous empirical evaluations of the ECHO model have demonstrated its efficacy, particularly in disseminating knowledge to providers on how to address rare healthcare conditions about which they had limited to no expertise. While it was originally developed to increase medical practitioners’ knowledge and self-efficacy in diagnosing and treating patients with rare physical health conditions such as Hepatitis C (Arora, 2015; Arora et al., 2014), it has been expanded for use with DD populations, such as ASD (e.g., Mazurek et al., 2017), and practitioners in educational and allied healthcare settings (Project ECHO, University of New Mexico School of Medicine, 2021). School-based personnel have reported increased knowledge and skills from participating in an ECHO network focused on promoting assistive technology use in schools (Root-Elledge et al., 2018). Given the multitude of advantages and low costs of the ECHO model, there is a need to explore the efficacy of this approach as a method for delivering parent education.

### Current Study Aims

To address the gap in accessible parent education programs for families of children with DD, this study evaluated the effects of participating in an innovative ECHO program for delivering free, accessible parent behavior management education and peer support to caregivers of children with DD. To date, there has been limited empirical evaluation of the efficacy of the ECHO model when applied with familial caregivers in lieu of professionals as the spokes. The purpose of this study was therefore to evaluate the efficacy of ECHO in improving caregivers’ knowledge, self-efficacy, confidence, and empowerment using a quasi-experimental, pre-post design in a statistically powered sample. The following primary hypotheses were tested: participating in the Caregiver ECHO program would increase caregivers’ knowledge of behavioral strategies for addressing CB, sense of empowerment to support children with DD, and their self-efficacy in managing CB. This study also explored whether participating would lead to decreased negative emotional reactions (sadness, anger, or anxiety) to CB as a secondary hypothesis. Finally, the ECHO model’s social validity with caregivers was assessed.

## Method

### Participants

Participants were the primary caregiver of a child with DD exhibiting CB at home during the COVID-19 pandemic. This study was conducted during the COVID-19 pandemic given the major losses of services that families of children with DD experienced, while recognizing that barriers to service access would persist even after the resumption of in-person services and education. Participants were recruited through social media posts, email listservs, pediatrics offices, online caregiver support groups, and word-of-mouth. Inclusion criteria were being the primary caregiver of a child who was: 1) enrolled in K-12^th^ grade, 2) had a DD diagnosis, 3) receiving or pursuing school-based special education services, 4) engaging in at least one form of CB (e.g., noncompliance, aggression, self-injury) at least once a week, and 5) not receiving other intervention supports for the CB for which they were pursuing ECHO support. Participants were recruited from across the USA, and the final sample consisted of caregivers from 10 states along the East Coast, West Coast, and in the Midwest.

An a priori power analysis using G*Power indicated that a minimum sample size of 28 would be needed to detect a medium effect of the intervention with a power of .80 (*α* = 0.05) for a Wilcoxon signed-rank test (Faul et al., 2007). Thirty caregivers (90% female, 10% male) participated in this study, each reporting on one child with DD. Most participants (83%) were White followed by Black (3%). Highest educational level achieved was widely distributed, with most having completed a bachelor’s degree (see Table 1). Fifty-three percent of participants lived in rural, 27% lived in urban, and 20% lived in suburban areas. Ninety percent of participants were biological and 10% were adoptive parents. No participants had previously participated in an ECHO program.

**Table 1.** Caregiver Participant (n = 30) Demographic Characteristics.

| Participant Characteristics | n(%) |
| --- | --- |
| *Race/Ethnicity* |  |
| Asian | 1(3) |
| Black | 1(3) |
| Hispanic/Latinx | 1(3) |
| Native Hawaiian/Pacific Islander | 0 |
| White | 25(83) |
| More than one | 2(7) |
| No response | 1(3) |
| *Highest education level* |  |
| High school graduate | 2(7) |
| Some college | 5(17) |
| Undergraduate degree | 1(3) |
| Master's degree | 11(37) |
| Doctoral degree | 3(10) |
| *Employment status before COVID-19* |  |
| Part-time | 7(23) |
| Full-time | 16(53) |
| Unemployed | 7(23) |
| Retired | 0(0) |
| Prefer not to say | 0(0) |
| *Employment change since COVID-19* |  |
| No change | 16(53) |
| Still employed, increased work hours | 6(2) |
| Still employed, but hours reduced due to personal/family obligations | 5(17) |
| Still employed, but hours reduced by employer | 5(17) |
| No longer employed | 5(17) |
| Furloughed | 5(17) |

### Measures

#### Child Demographic Measures

Caregivers completed a survey to provide demographic information on their child, specifically their child’s age, diagnoses, current educational supports, and support format (e.g., virtual, hybrid, in person). Caregivers were asked to report on their child’s community diagnoses (i.e., diagnoses conferred by a medical professional) of DD or a mental health disorder. To adhere to the ECHO model of keeping patients (in this case, children) deidentified from the hub team, diagnostic information was taken based on caregiver report alone.

#### Child Challenging Behavior Measure

Researchers developed a survey of CB frequency and severity, which participants completed before starting the ECHO program. Participants were asked to report on the types of CB displayed at home from a comprehensive pre-written list. For each topography of CB, definitions and examples were provided in parentheses (e.g., ripping, throwing, or swiping objects for property destruction). Frequencies were rated on a four-point Likert scale (1 = not currently, 2 = a few times a month, 3 = once a week, 4 = a few times a week, 5 = once a day, 6 = multiples times a day).

#### Behavioral Knowledge Test

Authors developed this measure, which included twenty multiple choice questions that assessed participants’ abilities to operationally define behavior, identify behavioral functions (e.g., attention, escape/avoidance), select appropriate intervention techniques based on behavioral function, define basic behavioral strategies (e.g., negative reinforcement), evidence-based strategies for measuring behavior change, and select intervention targets (Cooper et al., 2007). Four possible answers were given for each question. Tests were scored by calculating the total number of questions answered correctly. This assessment was completed at pre and post-test and differences between timepoints were calculated to assess knowledge change.

#### Difficult Behavior Self-Efficacy Scale

This self-report tool included five items rated from one to seven (not at all confident to very confident) to evaluate participants’ perceived confidence in their approaches to managing CB being effective (Hastings & Brown, 2002). The original instructions referenced CB displayed by a child with ASD – for the purpose of this study, instructions were modified to replace “autism” with “disability”. Independent psychometric evaluations of this tool in US and Korean samples confirmed good internal consistency (Cronbach’s *α* = .88) and excellent factor structure (Comparative Fit Index = 0.98)(Oh & Kozub, 2010).

#### Family Empowerment Scale (FES)

The FES was a 34-item questionnaire designed to measure caregivers’ self-reported empowerment to care for a child with disabilities (Koren et al., 1992). Each item was rated using a five-point Likert scale (0 = not true at all to 4 = very true). Originally developed as a two-subscale measure, psychometric analyses demonstrated substantial internal consistency (Cronbach’s *α* = .87 - .88), stable test-retest reliability (*r* = .77 - .85), substantial validity (overall *Kappa* = .77), and good factor structure. The FES is scored by summing all items to yield one overall score (Koren et al., 1992).

#### Emotional Reactions to Challenging Behavior Scale (ER-CB)

This self-report measure was completed at pre and post-test to evaluate the extent to which participants experienced negative emotional reactions to CB (Mitchell & Hastings, 1998). It included 23 items, each listing a different emotional reaction (e.g., shocked), and prompts respondents to rate how frequently they typically experienced each emotion when the person they were supporting demonstrated CB. Each item was rated using a four-point Likert scale ranging from “no, never” to “yes, very frequently”. Items were summed to yield two subscale scores: Depression/Anger (range = 0 – 30), and Fear/Anxiety (range = 0-15). The negative emotion subscales had good internal consistency and test-retest reliability and did not demonstrate susceptibility to social desirability response bias (Mitchell & Hastings, 1998).

#### Social Validity

A six-item measure of social validity was created for this study and administered at post-test. It surveyed participants on whether they believed that participating in the ECHO program increased their knowledge of behavior management strategies and ability to support children with DD, provided valuable access to expertise and a community of caregivers, increased their confidence in school-based special education supports by providing education around these services, and reinforced lessons learned through handouts that were disseminated at the end of each session. Items were rated using a five-point Likert scale (1 = Ineffective, 5 = Effective). The questionnaire also included an open-ended question following each quantitative question for participants to provide qualitative feedback.

### Procedure

This study had institutional review board approval. The Caregiver ECHO program was offered from October 2020 to May 2021to three cohorts in order to maintain small group sizes of 612 participants. Sessions were 1.25 hours, held in the evenings once a week for an eight-week period over Zoom® to accommodate families’ daily routines of dinner and bedtime. The hub team consisted of a session facilitator, a special educator, a clinical psychologist with expertise in DD, a school psychologist with 15 years of special education experience, a family navigator, and an ECHO coordinator who managed attendance and technical support. The special educator and clinical psychologist, who were doctoral-level board certified behavior analysts (BCBA-D) and had extensive experience in delivering parent education, presented all workshops. The family navigator and facilitator provided the lived experience perspective as the mothers of multiple children with DD.

Consistent with the original ECHO model, each session began with approximately five minutes of introductions, followed by a 20–25-minute workshop and a question-and-answer period. The remainder of the ECHO session was spent on one case presentation delivered by a participant. The eight-session series covered the following topics: 1) introduction to ECHO, reinforcement; 2) antecedent-behavior-consequence relationships of behavior; 3) antecedent-based intervention strategies, 4) understanding the functions of behavior; 5) consequence-based intervention strategies; 6) strategies for reducing CB and teaching replacement skills; 7) data collection; and 8) a review. A one-page workshop summary handout was provided after each workshop. The workshops were developed by the special educator on the hub team.

Each session, one participant presented on a CB their child was demonstrating for group-based problem solving. If participants endorsed observing multiple CBs at home at study enrollment, they were requested to choose the most troubling behavior as the focus of their case presentation. All participants were requested to complete a case submission form prior to the start of the ECHO program, in which they were asked to identify the most troubling CB their child was displaying and provide qualitative information on the child’s background information, strengths, what the targeted CB looked like and environment in which it occurred, previous strategies used to address the CB, and any additional pertinent information. Participants were asked to present each week on a voluntary basis. When the number of participants in a cohort exceeded opportunities to present, the ECHO facilitator compared cases to identify similar presentations and needs. One participant from subgroupings of similar presentations was invited to present, whose case discussion would likely yield greatest learning opportunities for the entire cohort. Presentations were 5-10 minutes long, followed by a period for clarifying questions from participants and the hub team. Presentations consisted of a single lecture slide that summarized content included in the case submission form. The ECHO facilitator used the presenting participant’s responses in the case submission form to prepare each lecture slide, asking for clarifying information when necessary.

As is consistent with the original ECHO model, during the remainder of the session, participants and hub members provided recommendations to the presenting participant on how to support the child’s positive behavior acquisition and reduce CB within home, educational, and family contexts. Participants were requested but not required to make recommendations, and each hub team member shared at least one recommendation from their respective discipline. Hub team members selected recommendations to share based on their expert judgement and the degree of relevance to the CB (e.g., prioritizing safety recommendations for dangerous self-injury) and from the perspective of their own disciplines to avoid providing overlapping advice with other hub team members. In the event where a participant provided a non-evidence-based recommendation (e.g., inaccurate explanation of special education law), a hub team member from the most relevant discipline used the example as a teachable moment to praise yet gently correct the participant’s recommendation. Following ECHO sessions, the hub team collaborated to create one written recommendations form that summarized all recommendations shared by the hub and spokes during the discussion, as well as additional recommendations from hub team members that were not shared during the discussion due to limited time. Participants were given access to all case recommendation forms as well as an online resources library that contained links to any resources referenced during sessions.

#### Modifications to the original ECHO model

The ECHO model is designed so that it can be flexibly applied across various audiences. Beyond the four core ingredients of the ECHO model (use of technology, best practice dissemination, case-based learning, and outcomes monitoring), the structure of an ECHO (e.g., duration, frequency, hub team structure) can be flexibly arranged while maintaining adherence to the original model (ECHO Institute, 2021). The primary modification in this application pertained to treating familial caregivers as spokes, whereas ECHO was originally designed for applications with professionals (e.g., physicians). Given that caregivers of children with DD regularly fill in for the roles of therapists, teachers, and aides, and that this was particularly the case due to the school and clinic closures during the COVID-19 pandemic, the ECHO Institute approved the application of the ECHO model with familial caregivers. A second minor modification to traditional ECHO procedures was that, due to the emotional strain of experiencing CB at home, the hub team regularly provided participants with encouragement and validation during their case presentations, above and beyond what is typically necessary within a professionally focused ECHO. A third modification was that full participation was incentivized with a $100 gift card to account for caregivers’ lost time and the expense of purchasing a Wi-Fi upgrade if needed. Full participation was defined as attending at least seven out of eight ECHO sessions and completing all pre and post-test measures. Traditionally, ECHO programs do not provide financial incentives to participants, given that participants receive the inherent benefit of free education and peer support. In this instance, the research team elected to provide financial incentives to help alleviate the burden of participation, given the increased strain caregivers were facing during the COVID-19 pandemic. Beyond this, all procedural components of the ECHO model remained consistent with the original approach.

##### ECHO Fidelity Ratings

As is recommended by the developers of ECHO, fidelity measures were collected for each session using a scorecard developed by the ECHO institute (Project ECHO, University of New Mexico School of Medicine, 2021). This scorecard assessed adherence by having respondents check whether each core component was observed. A study team member who did not participate in the ECHO program completed ratings for each session by watching session recordings. Across sessions, fidelity ratings averaged 100%.

#### Analysis Plan

Kolmogorov-Smirnov tests for homogeneity of variance demonstrated that the following dependent variable scores were non-normally distributed when separated out by timepoint: Behavioral Knowledge (post), ER-CB: Depression/Anger (pre, post), and ER-CB: Fear/Anxiety (post). As there were three cohorts of participants, and 22 out of 30 total participants presented a case during their ECHO participation, participants were compared on demographic variables, pre-test scores, and post-test scores based on their cohort and whether they presented a case. After confirming that there were no differences between cohorts nor groups based on these variables, hypothesis testing was performed using outcomes data for all 30 caregiver participants.

Descriptive analyses were performed to characterize participant demographics as well as their children’s demographics, diagnoses, and CB frequencies. To test main hypotheses, Wilcoxon signed-rank tests were calculated to determine whether the intervention resulted in significant improvement in dependent variables from pre-to post-ECHO. To account for multiple comparisons, a Bonferroni correction was applied to all *p*-values such that only values below *p* = 0.01 were considered statistically significant (*p* = 0.05/5). Treatment effect sizes (*r*) were calculated for all pre-post-test changes (Rosenthal, 1994).

## Results

### Child Demographics and Needs

Participants provided demographic information about their child for whom they were seeking ECHO support at study enrollment. Of 30 represented children, 100% had one DD diagnosis and 43% had multiple DD diagnoses. In addition to a DD diagnosis, 30% had one co-occurring mental health disorder and 33% had multiple co-occurring mental health disorders (see Table 2).

**Table 2.** Demographic Characteristics of Children from the Full Sample and the Subset of Children Presented for ECHO Network Consultation.

| Demographic Characteristic |  | % Presented Children<br>(n = 22) | % Total Children<br>(n = 30) |
| --- | --- | --- | --- |
| *Child Gender (M:F)* |  | 76:24 |  |
| *Child Diagnoses* | ADHD | 43 | 57 |
|  | Autism spectrum disorder | 38 | 40 |
|  | Anxiety Disorder | 24 | 57 |
|  | Behavioral disorders | 23 | 23 |
|  | Sensory Processing Disorder | 19 | ‡ |
|  | Genetic syndromes | 19 | ‡ |
|  | Depression or mood disorder | 10 | 17 |
|  | Learning Disability | 10 | 10 |
|  | Auditory Processing Disorder | 5 | ‡ |
|  | Intellectual Disability | 10 | 17 |
|  | Language Disorder | 5 | 13 |
|  | Other mental health or DD diagnosis | -- | 27 |

*Note.* ‡ Conditions were not surveyed as part of baseline assessment but were independently reported by participants during their case reviews

Participants additionally completed a survey at study enrollment to indicate all topographies of CB observed at home and their frequencies. The most frequent CB observed occurring multiple times per day were inattention (83%), hyperactivity (70%), and noncompliance (60%). Interfering stereotypical behavior (50%) and verbal aggression (43%) were also reported to occur multiple times per day in close to half of children. The least frequently endorsed CB was self-injury (3%). Greater detail regarding frequencies of CB across all children represented, as well as median frequencies, can be found in Table 3.

**Table 3.** Child Challenging Behavior Frequency at Pre-Test.

| Challenging Behavior | Median Response | Overall Frequencies (%) |  |  |  |  |  |
| --- | --- | --- | --- | --- | --- | --- | --- |
|  |  | Not<br>Currently | A Few<br>Times a<br>Month | Once a<br>Week | A Few<br>Times a<br>Week | Once a<br>Day | Multiples<br>Times<br>per Day |
| Hyperactivity | Multiples Times per Day | 10 | - | - | 7 | 13 | 70 |
| Inattention | Multiples Times per Day | 7 | - | - | - | 10 | 83 |
| Noncompliance | Multiples Times per Day | 3 | 3 | 3 | 10 | 20 | 60 |
| Verbal Aggression | Once a Day | 7 | - | 7 | 27 | 17 | 43 |
| Physical Aggression | A Few Times a Week | 17 | 13 | 13 | 23 | 7 | 27 |
| Property Destruction | A Few Times a Month | 30 | 20 | 13 | 20 | 10 | 7 |
| Self-Injury | Not Currently | 67 | 20 | 3 | 7 | - | 3 |
| Interfering<br>Stereotypies | Once a Day | 17 | 3 | - | 20 | 10 | 50 |
| Elopement | Not Currently | 53 | 17 | 3 | 17 | 7 | 3 |
| Other CB | Not Currently | 77 | 7 | - | 3 | - | 13 |

### Case Presentations

Twenty-two of the 30 represented children were described by their caregiver during a case presentation for feedback from the ECHO program. See Table 2 for demographic information of the subset of presented children (n=22) as well as all children represented (n=30). Presented children were 8.90 years old on average (SD = 4.15, range = 5-21) and primarily male (76%). Most common reported behaviors of concern for which participants requested case consultation were tantrums (19%), noncompliance with schoolwork or self-care (19%), anxiety (14%), aggression (14%), interfering restricted and repetitive behavior (14%), loud vocalizations (5%), frequent lying (5%), self-injurious behavior (5%), and difficulties transitioning (5%). Common diagnoses included disruptive disorders (i.e., oppositional defiant disorder, conduct disorder, impulse control disorder). Genetic disorders included Down and Duane syndromes. Children were enrolled in kindergarten through 12th grade at the time of participant enrollment in the study. Fifty-two percent of presented children were receiving special education services outlined by a formalized individualized education plan (IEP) at the time of the study, and 19% were receiving partial special education supports through a formalized 504 plan. Children were receiving schooling through general education primarily (52%), followed by special education (33%), home school (5%), private school (5%), or general education with a resource room (5%). Most children were attending school virtually (48%), followed by in-person four days per week (24%), in-person full-time (14%), and through a hybrid format (14%).

### Hypothesis Testing

Analyses of changes in scores from pre to post-test across all 30 participants supported all three main hypotheses: participating in the ECHO program had a very large effect on Behavioral Knowledge (*r=0.80*), FES scores (*r*=0.71), and Difficult Behavior Self-Efficacy scores (*r*=0.67). Concerning the secondary hypothesis, participating in the ECHO program resulted in significant decreases with large treatment effects on ER-CB: Depression/Anger (r=0.54). While significant improvements with medium effects were observed in ER-CB: Anxiety/Fear (*r*=0.46) scores, these improvements were not statistically significant at an alpha of 0.01, accounting for multiple comparisons (see Table 4).

**Table 4.** Effects of ECHO Participation on Main and Secondary Outcomes.

|  | Pre (n = 30) |  | Post (n = 30) |  | Z | p | r |
| --- | --- | --- | --- | --- | --- | --- | --- |
|  | Med | IQR | Med | IQR |  |  |  |
| Behavioral Knowledge | 14.50 | 12.00-16.00 | 16.50 | 15.75-18.00 | 4.41 | <.001* | 0.80 |
| Empowerment | 121.50 | 107.75-130.00 | 132.00 | 122.75-143.50 | 3.91 | <.001* | 0.71 |
| CB Self-Efficacy | 18.13 | 12.53-21.96 | 22.44 | 18.97-25.15 | 3.66 | <.001* | 0.67 |
| Depression/Anger | 13.00 | 9.00-19.00 | 9.50 | 6.75-12.25 | -2.98 | <.01* | 0.54 |
| Fear/Anxiety | 5.00 | 4.00-8.00 | 4.00 | 3.00-6.00 | -2.52 | <.05 | 0.46 |

Note: IQR = Interquartile range
r effect size ranges: $|r| < 0.1$ : no effect; $|r| = 0.1$ : small effect; $|r| = 0.3$ : medium effect; $|r| = 0.5$ : large effect\*p-value is significant using threshold of $p = 0.01$ (Bonferroni correction)

### Social Validity

On the whole, participants strongly agreed that participation in the ECHO program increased their knowledge of behavior management strategies and ability to support children with DD (M = 4.80, SD = 0.61), the ECHO model was effective in supporting children with DD (M = 4.73, SD = 0.58), the hub team provided expertise in supporting children with DD (M = 4.97, SD = 0.18), ECHO built a community of support for serving children with DD (M = 4.73, SD = 0.52), and that handouts effectively summarized each session’s workshop (M = 4.87, SD = 0.35). Zero participants provided “somewhat ineffective” or “ineffective” ratings for any of the questions.

Examining qualitative feedback, participants most frequently reported that access to a community of other caregivers decreased their sense of isolation (e.g., “I was so happy to find other parents like me. I feel like I am on an island by myself sometimes”), they learned about behavior management strategies (e.g., “I have learned so much from the hub and the network”), lessons learned from the interdisciplinary expert team and their peers was helpful (e.g., “It was encouraging and informative to hear the stories of others and to learn along with everyone. It felt like a positive community to be a part of”) and that they saw improvements in their child’s behavior after implementing recommended strategies (e.g., “We have implemented suggestions and seen changes”).

Participants also made recommendations for improving the program, specifically by keeping group sizes to eight or fewer to increase everyone’s opportunities to speak, opening the network to a mix of special education teachers and parents to build a larger community, including a self-advocate on the hub team with ASD or ADHD, and offering specific ECHO networks to families within the same communities (e.g., geographic area, racial/ethnic groups).

## Discussion

The goals of this study were to conduct a preliminary investigation of the efficacy of the ECHO model as an approach for delivering virtual parent education in behavior management to caregivers of children with DD. Results showed that the ECHO program was very effective for increasing caregivers’ knowledge of behavioral approaches for addressing CB, empowerment as caregivers of children with DD, and self-efficacy in responding effectively to CB. The program was also effective in reducing caregivers’ self-reported negative emotional reactions to CB. Finally, participants reported high satisfaction with and social validity of the ECHO model as an approach for delivering parent education around behavior management.

Increasing caregivers’ knowledge of behavioral strategies was one of the primary targeted outcomes of the ECHO program. While this study suggested that participating in an ECHO program may successfully increase behavioral knowledge, outcome measures did not capture whether ECHO participation changed caregivers’ knowledge of content targeted by the non-behavioral disciplines included in the hub team (school psychology, clinical psychology, and family navigation): parents’ rights within the school system and educational law, the process for pursuing an ASD or related evaluation, discussing psychotropic medication treatment options with physicians, parent resource and advocacy information, and self-care for caregivers. Future evaluations of the Caregiver ECHO program may consider evaluating the effects of participation on other areas of knowledge to fully capture potential positive impacts of the program.

Caregiver ECHO participation also showed important impacts on caregivers’ sense of empowerment, which is of note given that caregiver empowerment is less frequently a focus of parent education programs. Empowerment refers to “the process of becoming stronger and more confident, especially in controlling one’s life and claiming one’s rights” (Oxford University Press, 2023). Empowerment has been shown to be negatively impacted by family functioning (Wakimizu et al., 2017), and plays an important role in advocacy skills. Empowerment is less frequently an outcome targeted through parent education programs (Jackson et al., 2016), though it is an essential skill for parents of children with DD who often have to serve as advocates for their children to receive appropriate services and supports. The inclusion of a network of peers encouraging one another, along with an interdisciplinary hub team that provided guidance on special education rights, self-care, and resources, likely strengthened the impact of ECHO participation on empowerment. Given that participation in Caregiver ECHO was associated with very large effects on caregiver empowerment, using the ECHO model to disseminate support to caregivers may be a particularly effective and relevant approach to parent education.

Participation in the Caregiver ECHO program also was associated with to major increases in participants’ self-efficacy in managing CB. Caregiver self-efficacy has implications for children’s adjustment as well as overall parental competence and psychological functioning. Caregiver self-efficacy in managing CB has a great impact on daily life and functioning of both caregivers and children (Breitenstein et al., 2010; Jones & Prinz, 2005). Research has also shown that when caregivers feel confident and knowledgeable in implementing behavior strategies, fidelity of implementation increased (Casagrande & Ingersoll, 2017), which further translated into better treatment outcomes. Thus, increasing parenting self-efficacy is an essential target when providing education around behavior management strategies, and the Caregiver ECHO program successfully demonstrating increased self-efficacy suggests the value of continuing to make Caregiver ECHO programs available across communities.

Free or low-cost avenues for accessing parent education such as an ECHO program were particularly important for children with DD and their families who experienced service loss during the COVID-19 pandemic (Masi et al., 2021; Shorey et al., 2021). Even after the conclusion of the COVID-19 pandemic, continued access for families of children with DD remains imperative. Many children with DD have been shown to demonstrate high rates of co-occurring behavioral concerns (Emerson et al., 2014; Nicholls et al., 2020) that warrant access to behavioral therapies and supports. Yet, even beyond the COVID-10 pandemic, children with DD struggle to access these supports due to their geographic location, scheduling limitations, the deficit of qualified providers, or the costs of attending appointments as a function of copays, travel costs, babysitting fees, and lost work time (Vohra et al., 2014). It is therefore essential that effective, evidence-based approaches to parent education and support are developed that can be implemented for free or at low costs.

This study also showed that a novel adaptation of the ECHO model holds great promise for making a positive impact on families of children with DD by connecting them with peer emotional and social support. As was demonstrated by qualitative feedback, participants most frequently reported valuing access to a network of peers to reduce their social isolation and normalize their experiences as caregivers of neurodivergent children. While families of children with DD experienced significant anxiety, depression, and caregiver burden associated with the social isolation experienced during the COVID-19 pandemic (Iovino et al., 2021), caregivers also experience poorer mental health and social isolation outside of the pandemic (Peer & Hillman, 2014). Providing parent education in group formats that is easily accessible to families, such as virtually and at low-cost, is therefore essential for long-term family functioning and well-being.

Beyond testing the efficacy of the ECHO model in delivering parent education around other topics, more research is needed to evaluate how participation in ECHO networks directly impacts outcomes of participants and the students or children they represent (Hardesty et al., 2020). Future evaluation of the Caregiver ECHO program should capture direct and standardized measures of behavior change amongst children served. Further, additional randomized group design studies are needed to establish the causal relationships between participation in a Caregiver ECHO program and targeted outcomes. Future ECHO programs will also want to include strategic recruitment approaches to increase representation within their samples. A snowballing approach, for example, may be more effective in recruiting families from racial or ethnic minority backgrounds (Hughes et al., 1995). Finally, studies can investigate the extent to which the ECHO model builds capacity among other populations that support children with DD, such as special education teachers.

### Strengths and Limitations

The strengths of this study included having a sufficiently powered sample size to conduct hypothesis testing, the diversity of child behaviors and diagnoses represented in the sample, and the recruitment of families from across the United States representing homes from urban, suburban, and rural areas. Concerning limitations, as this study was designed as a preliminary efficacy study, a control group was not included. Further, while participants were surveyed on the frequency and topographies of CB their children were displaying at home at pre-test, a standardized measure of CB was not used at pre and post-test to facilitate a more objective measure of behavioral change. Participants were not surveyed on their age, and we therefore cannot speak to the diversity of parenting ability across the sample based on years of experience. There was limited racial and ethnic diversity across the sample. Nonetheless, the low-cost and accessible nature of the Caregiver ECHO program means that it can likely be used as a method for increasing equity in education and support to culturally and linguistically diverse families who may ordinarily struggle to access center-based parent education.

### Conclusion

This study demonstrated preliminary evidence that the ECHO model, which uses a hub-spoke framework to provide case consultation, education, and access to a network of peers, can serve as an inexpensive, effective approach for providing parent education around behavior management to caregivers of children with DD. The added benefits of increasing caregivers’ sense of empowerment and self-efficacy, as well as community building for caregivers, demonstrated the social validity of this approach for supporting families. It additionally added to the existing literature demonstrating the efficacy of delivering parent education virtually, which is significant for families of children with DD who experience high amounts of isolation due to the complexities of their child’s needs. Future research examining the impact of the ECHO model versus a comparison treatment on caregivers, such as a general parent education program, and that includes standardized measures of child behavioral change, would help elucidate the extent to which unique components of the ECHO model, such as peer case consultation and support, impacts caregiver and family outcomes above and beyond traditional parent education. Such discoveries would continue to drive the establishment of best-practice approaches to delivering parent education at low cost.

## Data Availability

All data produced in the present study are available upon reasonable request to the authors

